# Different approaches to quantify years of life lost from COVID-19

**DOI:** 10.1101/2021.05.13.21257193

**Authors:** Tamás Ferenci

## Abstract

The burden of an epidemic is often characterized by death counts, but this can be misleading as it fails to acknowledge the age of the deceased patients. Years of life lost is therefore widely used as a more relevant metric, however, such calculations in the context of COVID-19 are all biased upwards: patients dying from COVID-19 are typically multimorbid, having far worse life expectation than the general population. These questions are quantitatively investigated using a unique Hungarian dataset that contains individual patient level data on comorbidities for all COVID-19 deaths in the country. To account for the comorbidities of the patients, a parametric survival model using 11 important long-term conditions was used to estimate a more realistic years of life lost. As of 12 May, 2021, Hungary reported a total of 27,837 deaths from COVID-19 in patients above 50 years of age. The usual calculation indicates 10.5 years of life lost for each death, which decreases to 9.2 years per death after adjusting for 11 comorbidities. The expected number of years lost implied by the life table, reflecting the mortality of a developed country just before the pandemic is 11.1 years. The years of life lost due to COVID-19x in Hungary is therefore 12% or 1.3 years per death lower when accounting for the comorbidities and is below its expected value, but how this should be interpreted is still a matter of debate. Further research is warranted on how to optimally integrate this information into epidemiologic risk assessments during a pandemic.

## Manuscript

### Introduction

Quantifying the burden of the COVID-19 – or any other infectious disease – is not a straightforward undertaking, primarily due to the multifaceted nature of the problem: “burden” can be measured along several dimensions. Death, time spent with suffering (from anything from a mild cough to the need of invasive ventilation), onset of long-lasting sequelae, direct and indirect consequences of work absenteeism, utilization of the limited healthcare capacity can all be considered “burden” in some sense [1–6].

Perhaps the most widely used indicator of burden is the mortality associated with the epidemic [7]. This can be directly measured, it is considered to be very relevant and often correlated with other – more difficult to measure – indicators [8].

Simply calculating the number of deaths due to COVID-19 to measure the burden is associated with two inherent problems [9, 10]. The first is the definition of dying from the disease: how deaths are attributed in a multimorbid patient is not necessarily unambiguously defined, procedures might be different between countries or change over time. In addition to this uncertainty, deaths may be undercounted if patients are not tested for COVID-19 even post mortem. (This is the reason why the application of excess deaths, that is, the number of deaths minus the number of baseline value, i.e., the number of deaths without epidemic, estimated from historical data, is often called for [11]. Excess death calculation however relies on a baseline, a predicted value, which is always an extrapolation based on historical data, and as such, its reliability necessarily gets worse over time as we are further from the data from which it is predicted. In addition to this, excess death calculation cannot discern the – positive or negative – indirect effects from the direct effects of the epidemic.) These problems will not be addressed in the present paper.

The second problem, that will be investigated in detail is that raw deaths counts ignore the age of the deceased patient: this calculation gives equal weight to the death of a multimorbid, 80-year-old patient, who only had a few years of life expectancy even without the infection, to the death of a healthy 30-year-old patient, who had several decades of life expectancy. Thus, years of life lost (YLL), first used by Haenszel [12] is widely used to more accurately represent the burden of a disease [13–15].

YLL calculations either assume a fixed target age, to which years lost is measured, or – more typically – use a life table to calculate the expected remaining time (i.e., time lost) for each death [16].

For the COVID-19 pandemic, these calculations are however all biased upwards: patients dying from COVID-19 cannot be considered to be a random subsample from the general population. They are usually multimorbid, often having several long-term conditions [17, 18], which are themselves associated with reduced life expectancy. The correct calculation should take this into account, by subtracting the age at death from a lower expectation, resulting in a lower number of lost years of life. Calculation of the appropriate expectation is however not straightforward, as life table for patients with comorbidities – and especially for their arbitrary combinations – are not usually available, and are not feasible to produce.

A further important but often overlooked problem about YLL is the appropriate interpretation of its numerical value. “Losing a life year” sounds unequivocally negative, thus one is tempted to automatically assume that the ideal value of YLL is zero. This is not the case however. YLL is based on losing the remaining life expectancy, but this is never zero, so it is mathematically impossible to have a YLL of zero – even a patient dying at 110 years still loses some remaining life (1.46 years for males, 1.35 for females according to the Hungarian life table).

To tackle this problem, Marshall introduced the concept of life table norm for life years lost, which is the number of years lost by an “average” person in a given population. More precisely, YLL is calculated for a hypothetical cohort undergoing exactly the mortality specified by the life table and the resulting number of years lost is divided by the number of deaths [19, 20]. In this sense, the norm serves as a reference point, to which actual YLLs can be compared, as the norm signifies what loss is to be expected lacking any special mortality-modifying circumstance. Using a pre-pandemic life table, it can be used represent the expected YLL in individuals who die from causes other than the COVID-19 epidemic. How this should be integrated into the calculation (i.e., should it be simply deducted from the YLL, or only a fraction of it should be deducted) is a matter of debate.

The present study aims to quantitatively explore these issues for the COVID-19 pandemic using data from Hungary. Uniquely, individual patient level data on comorbidities is publicly available in Hungary for every reported death.

## Methods

Hungarian life table was obtained from the Human Mortality Database for 2017 (the latest available) [21]. Years of life lost for a deceased patient was defined as the expected time remaining according to the life table for the patient’s age and sex. YLL is then expressed either as “per death” (when total YLL is divided by the total number of deaths throughout the study period) or “per person-year” (when YLL is divided by the total person-year exposure of the whole background population throughout the study period).

Life table norm was calculated from this life table by summing the product of the number of deaths (from a fixed starting cohort with a size of 100,000) and expected remaining years for each age group and dividing it with 100,000.

COVID-19 deaths with individual patient level data on comorbidities were downloaded from the official Hungarian governmental COVID-19 website [22]. This gives information on each patient’s year-precision age, sex and comorbidities in an unstructured listing. Eleven comorbidities investigated by Hanlon et al [23] (atrial fibrillation, cancer, chronic obstructive pulmonary disease, dementia, diabetes, heart failure, hypertension, ischaemic heart disease, chronic renal failure, chronic liver disease, stroke) were identified using a regular expression based pattern matching. Search strings are given in Table 1.

**Table 1.**
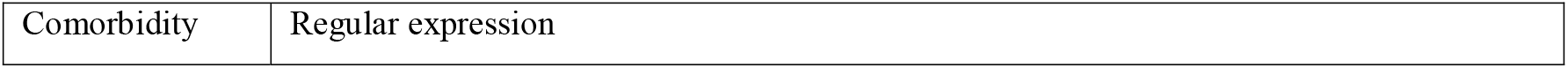
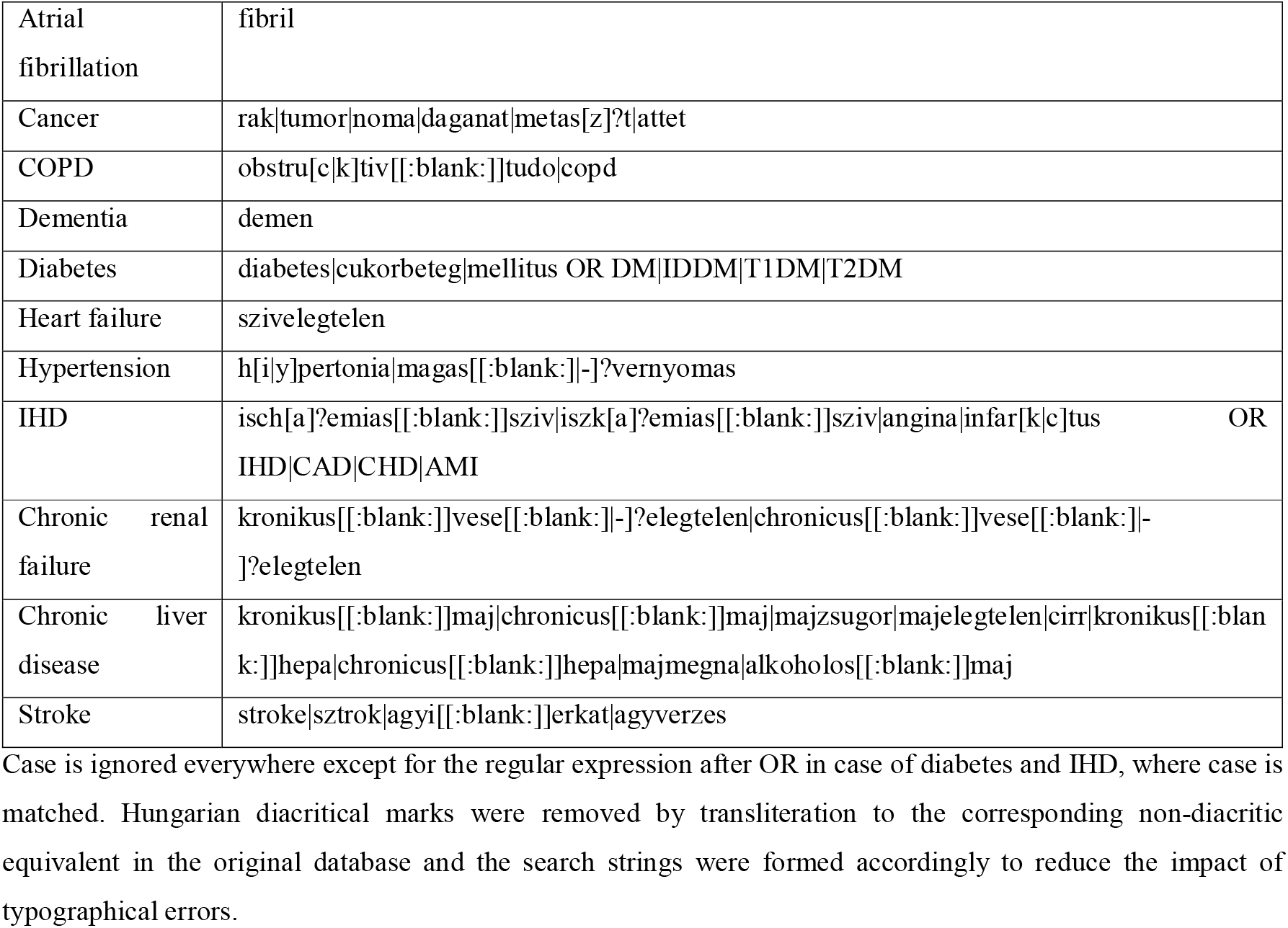
Regular expressions used for pattern matching comorbidities in the unstructured list of comorbidities.

Although it would be possible to decode any other comorbidity in the above fashion, these 11 were selected, as survival information were available for these from the literature. In more detail, survival models based on age and comorbidities were obtained from Hanlon et al [23] in the form of a parametric model using Gompertz distribution. Covariates were the 11 comorbidities and their interactions with age, and were assumed to govern the location parameter (i.e., rate) of the distribution [24]. Using the estimated parameters from Hanlon et al and each individual’s covariates, the survival curve for the particular patient was estimated, and expected survival time was calculated by integrating over a 0.1-year wide grid. Separate parameter sets were used for males and females. To account for the overall difference in the survival of the Hungarian population and the data – based on the population of Wales – from Hanlon et al, an offset correction was used: the ratio of the life expectancy from the intercept only model of Hanlon et al to the corresponding data from the Hungarian life table was used to multiply the life expectancy estimated from the survival model.

These survival models are only available for ages above 50 years, so the entire present analysis will be restricted to this age group.

Confidence interval for the prevalence estimates was calculated with Clopper-Pearson exact method [25]. Age-specific prevalence estimates were calculated using a spline-regression to smoothly model the effect of age without assuming any parametric functional form [26].

Calculations were carried out under the R statistical program package version 4.0.4 [27], using package flexsurv version 2.0 [24] and mgcv version 1.8-35 [28].

Full source code of the analysis script is available at https://github.com/tamas-ferenci/YLL_COVID19_Hungary.

## Results

As of 12 May, 2021, Hungary reported a total of 28,970 deaths from COVID-19, of which 27,837 occurred in patients above 50 years of age (13,667 females and 14,170 males). Figure 1 shows the distribution of the age of deaths according to sex. The size of the background population is 3,828,818 [29].

**Fig 1.**
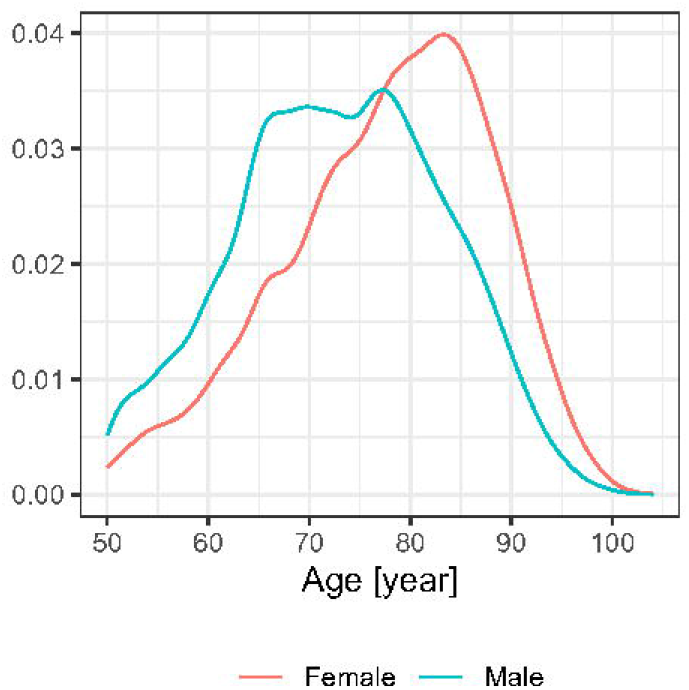
Distribution of the age of the deceased patients according to sex

Hungarians in this age group lost a total of 293,519 years of life due to COVID-19 using the ordinary life table approach. This means 10.5 years of life lost for each death (10.3 for females, 10.7 for males), and 0.064 years of life lost per person-year.

Prevalence of the 11 investigated comorbidities is shown on Table 2. In brief, the majority of the deceased patients had hypertension, around third of them were diabetic, and more than 10% had cancer or ischaemic heart disease. The prevalence of the remaining comorbidities was below 10%. Prevalence estimates by age and sex are shown on Figure 2.

**Table 2.** Prevalence of the 11 investigated comorbidities (95% confidence interval in parenthesis).

| Comorbidity | Prevalence [%] |
| --- | --- |
| Atrial fibrillation | 3.4 (3.2-3.6) |
| Cancer | 11.0 (10.6-11.3) |
| COPD | 3.9 (3.6-4.1) |
| Dementia | 7.6 (7.3-7.9) |
| Diabetes | 30.0 (29.4-30.5) |
| Heart failure | 7.5 (7.2-7.8) |
| Hypertension | 68.0 (67.4-68.5) |
| IHD | 14.6 (14.2-15.0) |
| Chronic liver disease | 0.8 (0.7-0.9) |
| Chronic renal failure | 5.4 (5.1-5.7) |
| Stroke | 2.9 (2.7-3.1) |

COPD: chronic obstructive pulmonary disease, IHD: ischaemic heart disease

**Fig 2.**
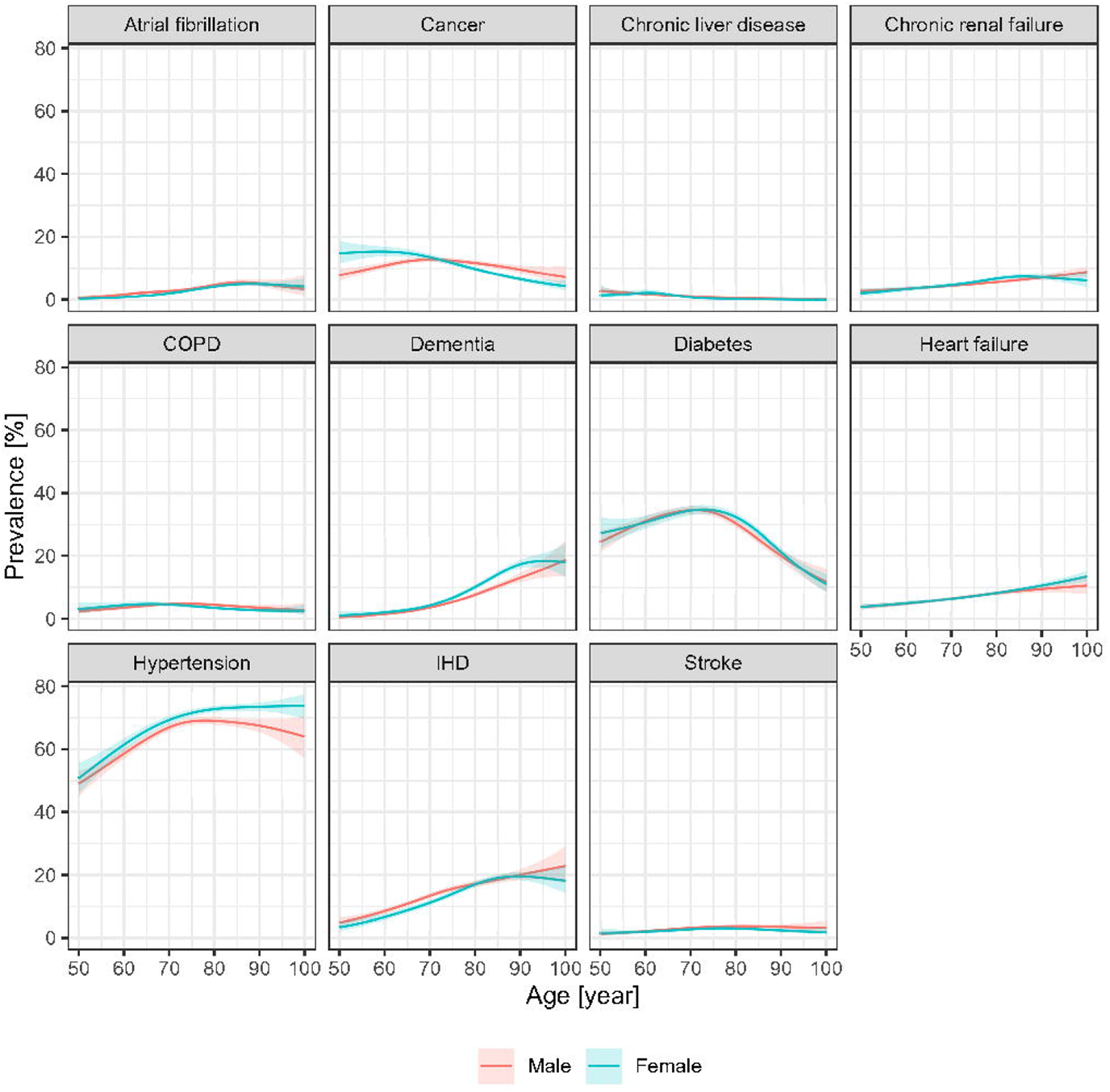
Prevalence of the investigated 11 comorbidities among the deceased patients according to age and sex with 95% confidence intervals; COPD: chronic obstructive pulmonary disease, IHD: ischaemic heart disease

Figure 3 depicts the distribution of the number of comorbidities by age and sex. Overall, 14.1% of the deceased patients had no comorbidity (from the investigated 11 ones), 36.4% had a single comorbidity, 33.7% had two comorbidities and 15.8% had more than two.

**Fig 3.**
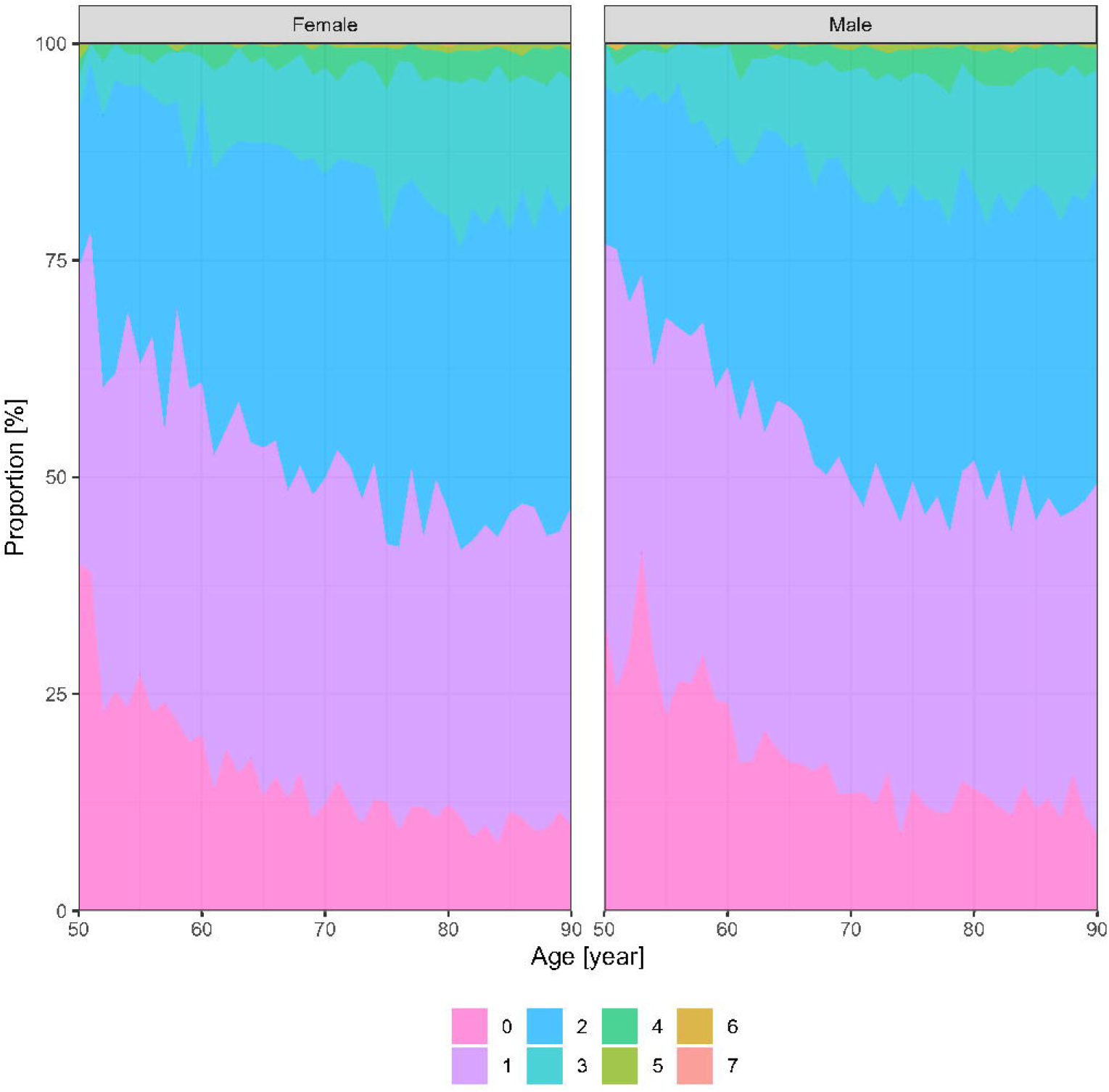
Distribution of the number of comorbidities of the deceased patients according the age and sex

Figure 4 shows the life expectancies of the life table contrasted with the ones provided by the survival model (at individual level, i.e., each dot represents a deceased patient). The difference is immediately obvious: when taking the comorbidities into account, the life expectancy is markedly lower when using the life table, which represents the general population.

**Fig 4.**
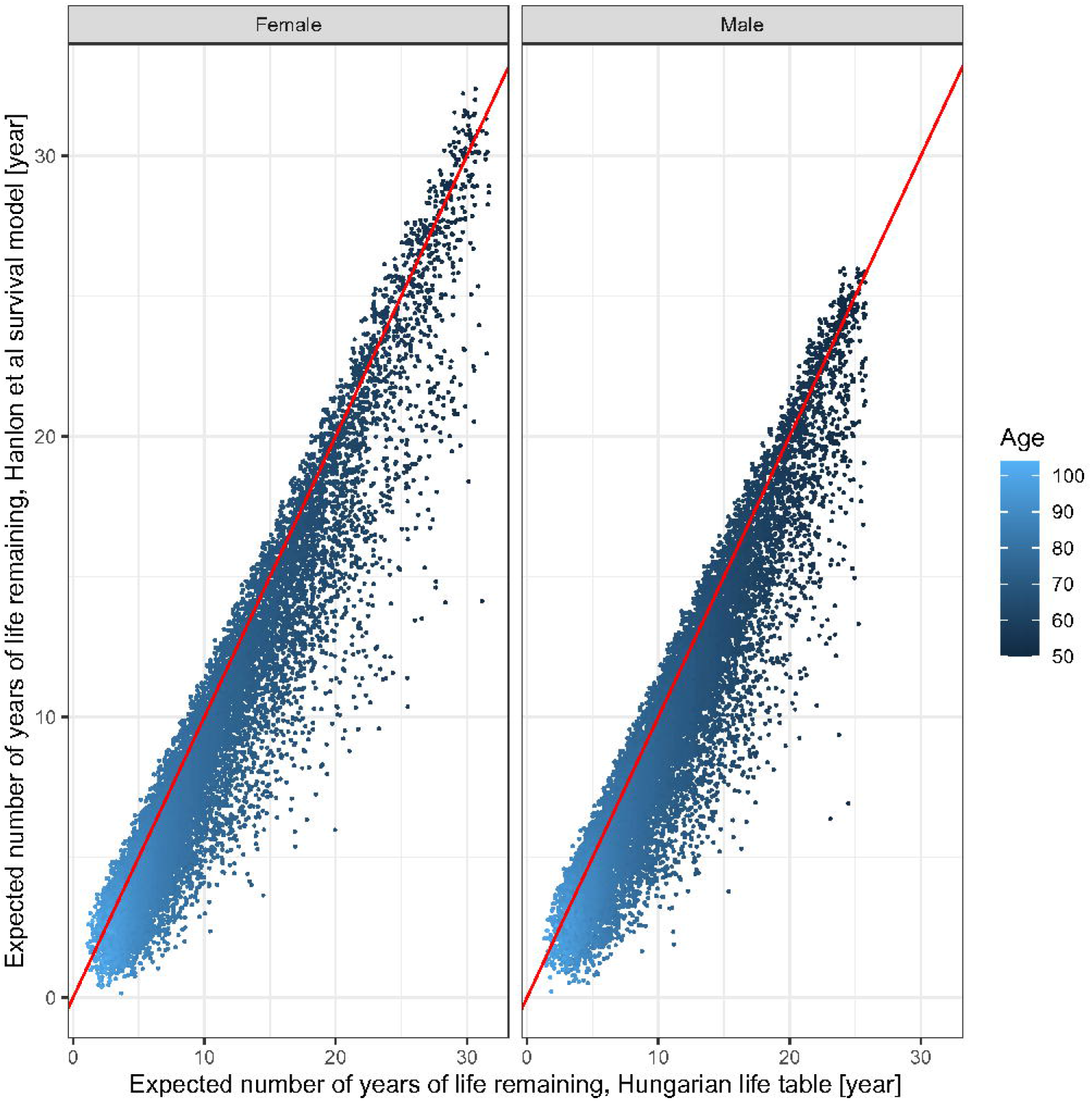
Jittered scatterplot of life expectancies from the Hungarian life table compared to the ones provided by the survival model, by age and sex; each dot represents a deceased patient

After adjustment for these comorbidities (i.e., using the survival model instead of the life table) the number of years of life lost decreases to 256,043 (9.2 YLL per death – 9.0 for females, 9.4 for males – or 0.056 YLL per person-year).

The life table norm of YLL according to the definition of Marshall is 11.1 years (10.3 years for females and 11.9 years for males). This was calculated from the pre-pandemic life table by summing the number of life years lost, the product of life expectancy and the number of deaths for each age group and then dividing it with the number of deaths, i.e., it represents the expected YLL per death due to all causes of death in Hungary, a developed Central European country, but the epidemic.

## Discussion

Burden of any disease can be characterized by its impact on the quality of life, and on life expectancy. The present study focuses on the latter question in the context of the COVID-19 epidemic.

As years of life lost better characterize the true burden of the disease than raw death counts, several studies assessed it for the COVID-19 pandemic. Aroles et al used the international COVerAGE-DB database to calculate YLL for 81 countries, and report a total of 20.5 million years lost as of 6 January, 2021 [30]. They use both reported deaths and excess deaths to account for the potential undercounting. In line with the message of the present paper and that of Hanlon et al, they also note that this number is likely highly biased upwards, thus emphasize the usage of YLL ratios (when YLL is compared to that of other causes, such as influenza, where a similar impact of multimorbidity can be expected). Quast et al used the ordinary life table approach in the United States and report 1.2 million years of life lost to COVID-19 as of 11 July 2020 [31]. In this study the authors accounted for the effect of multimorbidity by simply reducing the life table remaining years by 25%. Mitra el al evaluated the years of life lost in the United States, Italy and Germany using a fixed target age instead of the life table approach [32]. Rommel et al reported 305,641 years of life lost in Germany in 2020 using the usual life table method [33]. Goldstein and Lee estimated 11.7 YLL per death, but they make no attempt to account for the comorbidities of the deceased patients [34].

The paper of Hanlon et al was the first attempt to quantitatively investigate how the accounting for the comorbidities of the patient impacts the estimated years of life lost [23]. However, they had no access to individual patient data on comorbidities and therefore had to rely on an approximate reconstruction of the individual data from aggregate data which is necessarily less efficient and reliable.

Briggs et al performed a similar analysis, but using an aggregate indicator instead of actual, individual-level data for adjusting for the comorbidities [35].

A unique characteristic of the Hungarian data is that it allows public access to detailed, individual level comorbidities for every registered COVID-19 deaths (>27,000). This allowed a more direct analysis.

Indeed, accounting for 11 comorbidities identified from the Hungarian database decreased the years of life lost by about 12%, which is the first such direct result in the literature to our best knowledge.

The life years lost are however below the norm (as defined by Marshall) even without taking the effect of comorbidities into account. This result might seem counterintuitive at first glance, but is actually entirely possible. Consider the example of Rubo el al [36]: the “public health burden” of a serial killer specifically targeting people above 90 years of age measured in YLL would be almost surely below the norm due to the age of the victims. Simply comparing the YLL to the norm would therefore, at face value, indicate that the activity of a serial killer is beneficial to the public health. This warns us that how the norm should be integrated into the evaluation of the actual YLL of a specific cause is a complicated question.

A few things should be noted about this approach. First, a national life table was used for the ordinary YLL calculations, instead of using a fixed value, or subnational tables [37]. Second, the whole period was used, i.e. no attempt was made to separate different phases of the epidemic. This might be relevant, as different population groups could be affected due to changing non-pharmaceutical interventions, or the introduction of a new variant might alter the age-dependent risk for the same group.

The major limitation of the present study is the application of an external survival model to calculate the potential life expectancy for the patients who died. A logical and important research step would be the calculation and application of a survival model for the comorbidities from the same Hungarian population. Administrative/financial data has been extracted and successfully applied from the Hungarian healthcare system’s databases for biomedical research [38–40], so this endeavour seems to be feasible in the future.

Perhaps the most important strength of the present is study is the application of the detailed, individual-level comorbidity database (available for more than 27,000 deaths). However, the data quality is poor, comorbid diseases are entered without any form of standardization, with many typographical errors, arbitrary usage of Latin and Hungarian terminologies, arbitrary usage of abbreviations etc. We tried to overcome these limitations by using carefully selected search expressions to identify the comorbidities, but no formal analysis on the sensitivity or specificity was carried out. We also have no systematic validation on the correctness of the recorded comorbidity data.

A third limitation is that only 11 comorbidities were used and no information was available on the severity of the comorbidity.

A final limitation is the application of YLL itself. A recent paper of Rubo et al argues that the application of YLL is incorrect when there is no clear, causal model of mortality [36]. While strictly speaking we almost never have a perfect such model, this is much less of a problem when the age of death is markedly lower than the age of death in the general population, as this implies that the deaths can be strongly attributed to the investigated factor (as other are unlikely to cause death at early age). Of note, Rubo et al approvingly refer to the norm-adjustment of Marshall, but take a middle ground between not using it at all, and subtracting the entirety of the norm from the calculated YLL, arguing that this should be decided on how mono-causal is the attribution between the investigated factor and the death. (That is, no subtraction should be made if there is a purely mono-causal relationship, and 100% of the norm should be deducted if there is no plausible causal mechanism.) What fraction should be deducted in a situation like the COVID-19 epidemic, and whether the determination of this fraction is a feasible task at all is perhaps the most intriguing future research direction.

Finally, YLL focuses only on mortality, neglecting the quality of life (QoL) aspect. This is important in two, opposite directions. First, COVID-19 sometimes causes long-lasting sequelae which are detrimental to QoL [41, 42], thus a more complete analysis should also consider this, even if the patient survived the disease. Another, and likely more important consideration is that many of the – typically multimorbid – patients dying from COVID-19 very likely had a reduced QoL even before the infection, so an analysis that adjusts for QoL will reveal an even fewer number of – quality-adjusted – life years lost.

## Conclusion

Evaluation of the years of life lost is crucial, as it provides a much more relevant insight into the burden of the epidemic than raw death counts. The actual calculation however is not straightforward, and depends on many assumptions which should be carefully assessed. Further research is warranted on how to optimally integrate this information into epidemiologic risk assessments during a pandemic.

## Data Availability

Availability of data and material (data transparency): https://github.com/tamas-ferenci/YLL_COVID19_Hungary

## Acknowledgement

The author would like to express his sincere gratitude to Viktor Müller (Eötvös Loránd University) for his suggestions.

## Declarations

### Availability of data and material (data transparency)

https://github.com/tamas-ferenci/YLL_COVID19_Hungary

### Code availability (software application or custom code)

https://github.com/tamas-ferenci/YLL_COVID19_Hungary

## Authors’ contributions

TF planned the research, carried out the analysis and prepared the manuscript.

## Ethics approval

Not applicable.

## Consent to participate

Not applicable.

## Consent for publication

Not applicable.

